# SARS-CoV-2 genomes from Oklahoma, USA

**DOI:** 10.1101/2020.09.15.20195420

**Authors:** Sai Narayanan, John C Ritchey, Girish Patil, Teluguakula Narasaraju, Sunil More, Jerry Malayer, Jeremiah Saliki, Anil Kaul, Akhilesh Ramachandran

## Abstract

Genomic sequencing has played a major role in understanding the pathogenicity of severe acute respiratory syndrome coronavirus 2 (SARS-CoV-2). With the current pandemic, it is essential that SARS-CoV-2 viruses are sequenced regularly to determine mutations and genomic modifications in different geographical locations. In this study we sequenced SARS-CoV-2 from 5 clinical samples obtained in Oklahoma, USA during different time points of pandemic presence in the state. One sample from the initial days of the pandemic in the state and 4 during the peak in Oklahoma were sequenced. Previously reported mutations including D614G in S gene, P4715L in ORF1ab, S194L, R203K and G204R in N gene were identified in the genomes sequenced in this study. Possible novel mutations were also detected such as G1167V in S gene, A6269S and P3371S in ORF1ab, T28I in ORF7b, G96R in ORF8. Phylogenetic analysis of the genomes showed similarity to viruses from across the globe. These novel mutations and phylogenetic analysis emphasize the contagious nature of the virus.

## Introduction

Towards the end of 2019, several individuals with signs of pneumonia reported to hospitals in Wuhan, the capital of Hubei province in Central China. The etiological agent was identified to be a novel coronavirus (SARS-CoV-2/ nCoV-19) on 7^th^ January 2020(1). Human to human transmission was recorded around the same time(2). By the end of January 2020, WHO declared a ‘public health emergency of international concern’(3). As of September 5^th,^ 2020, a total of 26,696,160 confirmed patients and 875,943 deaths have been reported worldwide and 63,187 registered cases and over 850 deaths in the state of Oklahoma, USA(4).

Numerous coronaviruses infecting different animal species including humans have been identified. SARS-CoV-2 is an enveloped positive sense single stranded RNA virus belonging to the genus *Betacoronaviru*s (subgenus *Sarbecovirus*) in the *Coronaviridae* family (order: *Nidovirales*). Based on genomic sequence analysis, it is reported to have originated from bats(5) and pangolins(6, 7). Other previously identified coronaviruses known to infect humans include SARS-CoV-1, MERS-CoV, HCoV-NL63, HCoV-229E, HCoV-OC43 and HCoV-HKU1(8–13).

Genomic sequence data is important in identifying, characterizing and understanding pathogens (14, 15). It can shed light on pathogenicity, virulence, drug/vaccine targets, mutation sites etc. and can also be critical in source attribution and determining microbial provenance (16). The first genome sequence of SARS-CoV-2 was made available on January 10^th^ (GenBank ID: MN908947.3) (17). Multiple genomic sequences of SARS-CoV-2 from all over the world have since been deposited in public data bases such as GenBank and GISAID (Global Initiative on Sharing All Influenza Data; www.epicov.org) (18). This has facilitated extensive genomic studies leading to the identification of several mutations in the genome that can influence infectivity and virulence of the virus (16, 19, 20).

The genome of SARS-CoV-2 is similar to many other pathogenic coronaviruses and has multiple genes that code for different proteins such as S gene (Surface glycoprotein), N gene (nucleocapsid phosphoproteins), M gene (membrane glycoprotein), E gene (envelope protein), and open reading frames, such as ORF1a, ORF1b ORF3a,ORF6, ORF7a, ORF7b, ORF8 and ORF10 (16, 21). The leading sequence of the viral genome is the sequence for ORF1ab, which encodes for multiple proteins including replicase polyproteins, non-structural proteins, papain-like proteinase, RNA dependent RNA polymerase etc., which are essential for replication and survival in the host (21). Though the exact function of ORF3a is yet to be clearly understood, it is believed to play a major role in viral release after replication in SARS-CoV-1 (22). ORF7 and ORF8 code for accessory proteins, the functions of which are yet to be clearly understood (23, 24). Minimal roles of ORF7 and 8 in viral replication have been reported in SARS-CoV-1 alongside apoptosis stimulation of host cells(25).

The huge repository of sequence data in open-source databases such as the GenBank-NCBI and GISAID has facilitated the identification of numerous mutations and single nucleotide polymorphisms (SNPs) in the SARS-CoV-2 genome. SNPs in the genome that result in commonly reported non-synonymous mutations such as P4715L in ORF1ab, D614G in S gene, R203K and G204R in N gene are some of the commonly reported. P4715L in ORF1ab is believed to play a major role in interaction with other proteins that regulate RNA dependent RNA polymerase (26). D614G (Aspartate to Glycine) mutation in the S gene has been reported to result in increased transduction into human epithelial cells (20). N gene mutations R203K and G204R are believed to increase viral fitness, survival and adaptation to humans (27).

In this study, we sequenced the genome of SARS-CoV-2 from 5 human clinical samples received at the Oklahoma Animal Disease Diagnostic Laboratory (OADDL) at various times during the SARS-CoV-2 pandemic in Oklahoma. Multiple mutations were detected in the genomic sequences including those already reported as well as previously unreported mutations.

## Materials and methods

This study was approved by the Institutional Review Board (Application number: IRB-20-357) at Oklahoma State University, Stillwater OK 74078, USA.

### Clinical samples and processing

Five Nasopharyngeal swabs collected from human patients received at OADDL for COVID-19 testing between March 2020 – July 2020 were used in this study. Nucleic acid extraction was performed using MagMax Viral Pathogen Nucleic acid isolation kit (Thermofisher, MA, USA) as per manufacturer’s recommended protocols. Viral presence was detected by real-time PCR using TaqPath COVID-19 Multiplex Diagnostic Solutions (Thermofisher, MA, USA). All samples had a cycle threshold value between 18-22.

### Genomic sequencing

cDNA was synthesized from extracted RNA from five clinical samples that were positive for SARS-CoV-2. cDNA was then PCR amplified using ARTIC V3 primers (https://github.com/artic-network/artic-ncov2019/tree/master/primer_schemes/nCoV-2019/V3) to obtain overlapping segments of the whole viral genome. DNA library repair (SQK-LSK-109, Oxford Nanopore Technologies, UK), Solid Phase Reversible Immobilization (SPRI) paramagnetic beads clean-up (AMPure XP, Beckmann Coulter, CA, USA) and adapter ligation and barcoding (NBD-001, Oxford Nanopore Technologies, UK) were done as per manufacturer recommendations. Libraries were then pooled and sequenced using MinION (Oxford Nanopore Technologies, UK) platform following manufacturer recommendations.

### Genome Assembly, Alignment and Phylogenetic analysis

Sequences obtained were assembled *de novo* using Canu (28). To further obtain a reliable consensus genome assembly, *de novo* assemblies and sequence output files were assembled to the SARS-CoV-2 reference genome Wuhan Hu-1 (GenBank ID: MN908947.3) with minimap2 (29) and Nanopolish (30).

To assess uniqueness of the genomes sequenced in this study, MAFFT (31, 32) was used to align whole sequences of the 5 genomes to SARS-CoV-2 reference genome (NC_045512.2).

Gene predictions on the consensus assemblies were made using Viral Genome ORF Reader 4 (33) (VIGOR4) using a curated library available in the Virus Pathogen Resource (ViPR) (34) database. Individual genes were aligned to SARS-CoV-2 Wuhan Hu-1 genome from NCBI (GenBank ID: MN908947.3) using MUSCLE aligner in MEGA-X (35) to identify SNPs and changes in the amino acid produced by the gene.

To assess similarity to previously reported genomes, a phylogenetic analysis was made using 9072 genomes of SARS-CoV-2 from the GenBank database and the 5 viruses sequenced in the study. A General Time Reversible (GTR) substitution model based Unweighted Pair Group Method with Arithmetic Mean (UPGMA) alignment was constructed using MAFFT and FastTree (36). Clade definitions for the sequences were identified using 9 marker variants reported for classification in the GISAID database (18).

## Results and Discussion

Five clinical rRT-PCR positive samples from different periods of the pandemic in Oklahoma, USA were chosen for the study. One of the samples (Oklahoma-ADDL-1) was received during the initial stages (April 2020) of the pandemic. An increased incidence rate of the disease was observed by the end of May 2020 (4). Three of the samples (Oklahoma-ADDL-2,3,4) sequenced were received during this period and the last sample (Oklahoma-ADDL-5) was received one month (June 2020) after this period. More than 4000X coverage was obtained at the end of sequencing for all the samples. Following *de novo* assembly with Canu, consensus genomes were obtained after reference genome assembly with nanopolish and minimap2.

The five viral genome assemblies were aligned to the reference genome (SARS-CoV-2 Wuhan-Hu-1 NC_045512.2) using MAFFT and genome similarities to the reference isolate were calculated (Table 1). Most of the genomes showed more than 99% similarity. For Oklahoma-ADDL-5, only a partial sequence could be generated and hence showed a lower 86.836% similarity. This could be due to reduced amplification during amplicon generation and also due to reference assembly. Other than Oklahoma-ADDL-5, Oklahoma-ADDL-1 sequenced from a clinical sample obtained during the beginning of the pandemic in Oklahoma (April 2020) showed a lower similarity to the reference genome when compared to the other four isolates.

**Table 1:**
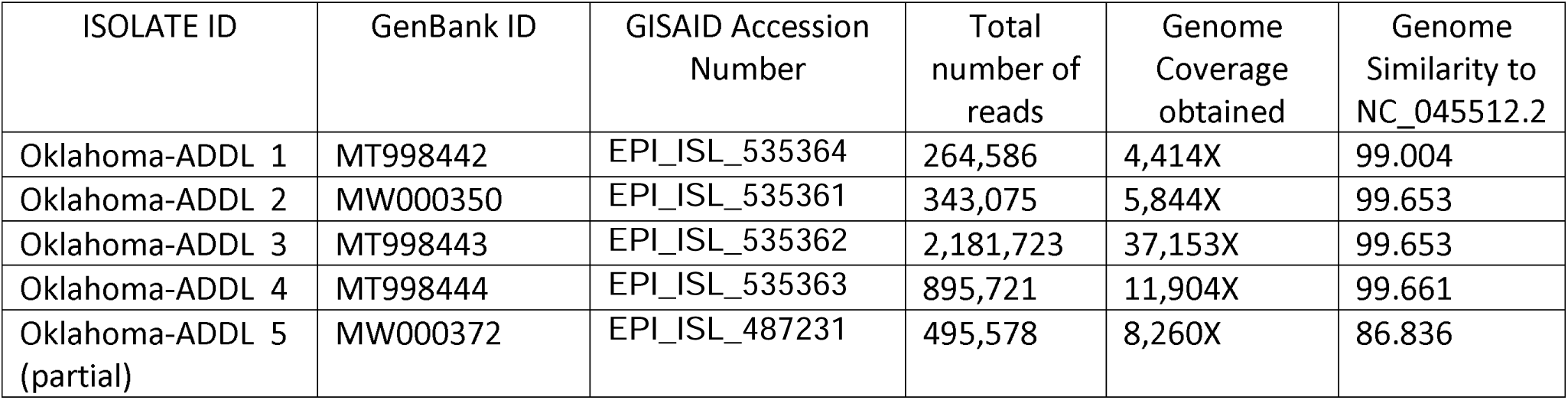
Genome similarity of 5 sequenced genomes when compared to NCBI reference genome (NC_045512.2) along with total reads generated and genome coverage obtained

| ISOLATE ID | GenBank ID | GISAID Accession Number | Total number of reads | Genome Coverage obtained | Genome Similarity to NC_045512.2 |
| --- | --- | --- | --- | --- | --- |
| Oklahoma-ADDL 1 | MT998442 | EPI_ISL_535364 | 264,586 | 4,414X | 99.004 |
| Oklahoma-ADDL 2 | MW000350 | EPI_ISL_535361 | 343,075 | 5,844X | 99.653 |
| Oklahoma-ADDL 3 | MT998443 | EPI_ISL_535362 | 2,181,723 | 37,153X | 99.653 |
| Oklahoma-ADDL 4 | MT998444 | EPI_ISL_535363 | 895,721 | 11,904X | 99.661 |
| Oklahoma-ADDL 5 (partial) | MW000372 | EPI_ISL_487231 | 495,578 | 8,260X | 86.836 |

Genes were predicted using VIGOR4 and individual genes were aligned to their respective NCBI reference genes using MUSCLE aligner with UPGMA alignment in MEGA-X to assess mutations in the genome. Using MEGA-X visualization tool, various silent and missense mutations were detected. The missense/non-synonymous mutations detected in major genes are listed in Table2. While non-synonymous mutations were detected in ORF1ab, ORF1a, S, N, ORF3a, ORF7, ORF8, none were detected in Envelope (E) gene, Membrane glycoprotein (M) gene or ORF10 gene.

Non-synonymous mutation at amino acid location 81 (C→T; Arginine → Cysteine), which codes for nsp2 was present in all the genomes sequenced in the study. The potential implication of this mutation is unknown. nsp2 along with nsp3 are known to play major roles in pathogenesis (37) of SARS-CoV-2 in humans. A few other possibly novel and previously reported non-synonymous mutations in ORF1a and ORF1ab were also identified. A previously reported mutation in the ORF1ab gene P4715L(38) (Proline to Leucine) was recorded alongside novel mutations at various amino acid locations. P4715L mutation has been implicated to play a major role in interaction with other proteins that regulate RNA Dependent RNA polymerase activity. P3371S (Proline to Serine) mutation in ORF1ab and ORF1a was detected in Oklahoma-ADDL-5. Oklahoma-ADDL-4 carried mutation T4412A (Threonine to Alanine) in ORF1ab, while Oklahoma-ADDL-2 and 3 carried mutation A6269S (Alanine to Serine). ORF1ab has multiple functions including RNA dependent RNA polymerase activity, helicase activity, Fe-S cluster binding, Zn^-^ binding activity, methyltransferase activity(39) etc. Functional implications of these mutations are still unknown and further studies to understand functional changes caused by these mutations may aid in better understanding the pathogenesis of the viral isolates found in Oklahoma.

Non-synonymous mutations were also found in S, ORF3a, ORF7b, ORF8 and N gene (Table 2). A previously reported mutation in S gene - D614G(40) (Aspartate to Glycine), was identified in all the genomes sequenced. The D614G mutation has been reported to cause a decrease in PCR cycle thresholds, suggestive of higher upper respiratory tract viral load (41, 42) in the host. A previously reported deleterious variation in the protein expressed from ORF3a, Q57H(40, 43), was recorded in Oklahoma-ADDL-1, the genome isolated at the beginning of the pandemic in Oklahoma. This mutation was not recorded in genomes isolated at other times in the state. The N gene also carried other previously reported mutations (Table 2). Oklahoma-ADDL-4, carried a non-synonymous mutation S194L(19) while Oklahoma-ADDL 2,3,5 carried R203K and G204R(40).

**Table 2:**
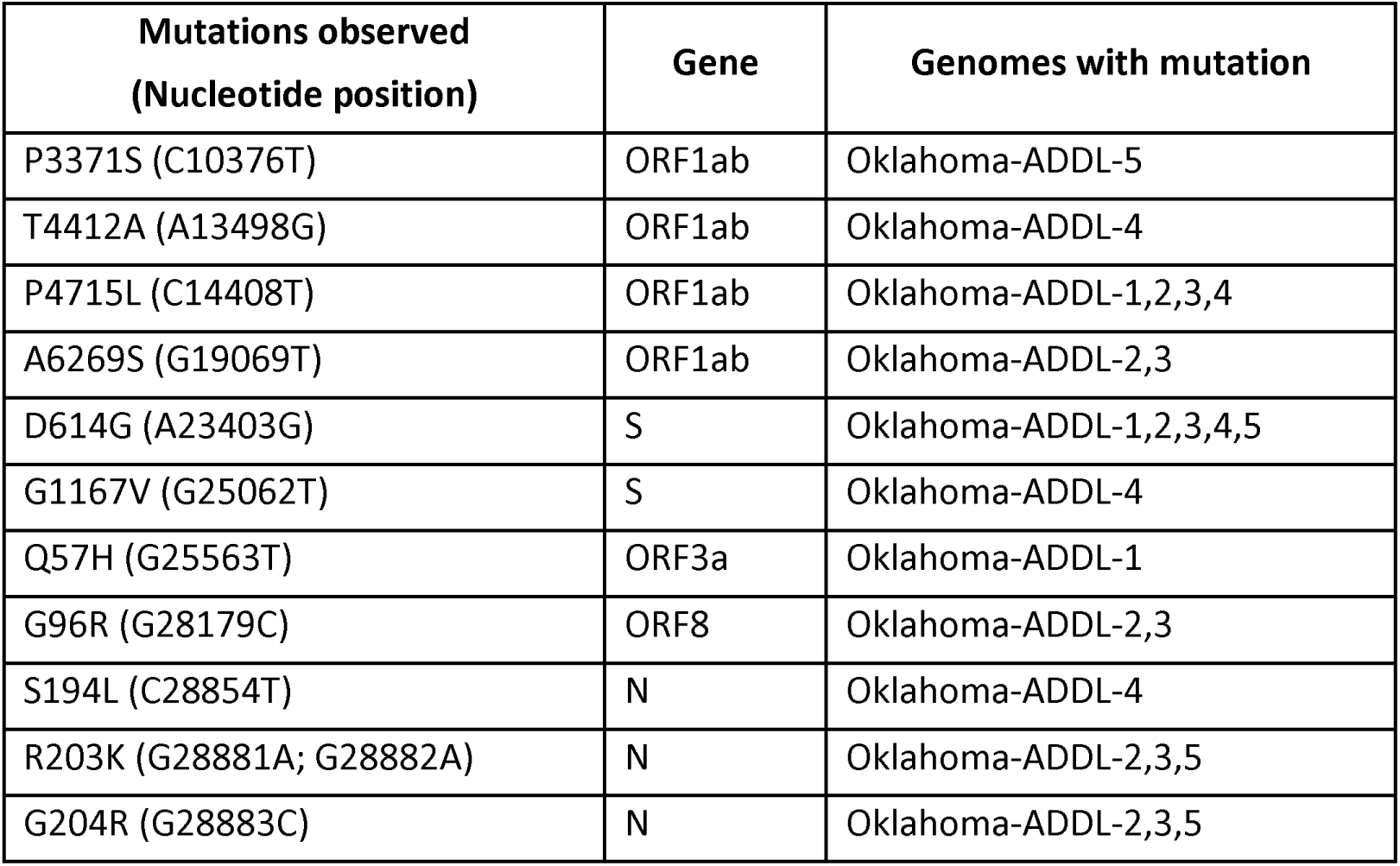
Non-synonymous mutations detected and their respective amino acid changes when compared to NCBI reference genome (SARS-CoV-2, Wuhan Hu-1, NC 045512.2).

| **Mutations observed**<br>**(Nucleotide position)** | **Gene** | **Genomes with mutation** |
| --- | --- | --- |
| P3371S (C10376T) | ORF1ab | Oklahoma-ADDL-5 |
| T4412A (A13498G) | ORF1ab | Oklahoma-ADDL-4 |
| P4715L (C14408T) | ORF1ab | Oklahoma-ADDL-1,2,3,4 |
| A6269S (G19069T) | ORF1ab | Oklahoma-ADDL-2,3 |
| D614G (A23403G) | S | Oklahoma-ADDL-1,2,3,4,5 |
| G1167V (G25062T) | S | Oklahoma-ADDL-4 |
| Q57H (G25563T) | ORF3a | Oklahoma-ADDL-1 |
| G96R (G28179C) | ORF8 | Oklahoma-ADDL-2,3 |
| S194L (C28854T) | N | Oklahoma-ADDL-4 |
| R203K (G28881A; G28882A) | N | Oklahoma-ADDL-2,3,5 |
| G204R (G28883C) | N | Oklahoma-ADDL-2,3,5 |

A few possible novel mutations were also identified in S, ORF3a, ORF7b, ORF8 and N genes (Table 2). A mutation in the S gene, G1167V (Glycine to Valine) was identified in Oklahoma-ADDL-4. T28I in ORF7b gene, a non-synonymous mutation (Threonine to Isoleucine) in the genome Oklahoma-ADDL-5 and in ORF8, G96R non-synonymous mutations resulting in Glycine to Arginine were noted in Oklahoma-ADDL - 2 and Oklahoma-ADDL-3. These mutations in the genome indicate presence of multiple variations of the virus in Oklahoma.

A phylogenetic tree was constructed using 9072 genomes available in the NCBI repository. The nearest neighbors to the genomes were found to be isolates reported from San Diego and Atlanta in USA and from Greece and Australia. While the Oklahoma-ADDL-1 and 5 were phylogenetically more related to isolates reported from Australia, Oklahoma-ADDL-2 and 3 were phylogenetically related to isolates reported from Greece and Atlanta, USA. Oklahoma-ADDL-4 was phylogenetically related to the isolate reported from San Diego USA. Oklahoma-ADDL-1 was identified to be in clade-GH and Oklahoma-ADDL-2,3,4,5 were identified to be in clade GR as per annotations of different marker variants in the GISAID database. The presence of multiple isolates within the Oklahoma sheds light on contagious nature of these viruses.

The genomes sequenced have been submitted to GISAID and GenBank (Table 1)

## Conclusion

We determined the sequence five SARS-COV-2 genomes, collected between March to July from Oklahoma USA, using MinION by Oxford Nanopore Technologies. Genome assembly and annotation studies identified several new mutations as well as previously reported mutations. Notably, presence of D614G mutation S gene was found in all the isolates, thus indicating presence of highly virulent strains in Oklahoma. Detection of multiple mutations in the viral genomes collected from a narrow geographic region within a few months of the Pandemic underscores the ability of SARS-CoV-2 to undergo rapid genomic alterations that can potentially influence host susceptibility, pathogenicity and virulence. Phylogenetic analysis of the virus genomes revealed high similarities in gene sequences with isolates reported from Australia, Greece, Atlanta and San Diego in USA, indicating origin of different isolates arising from multiple introductions in the state. Further mass sequencing of clinical isolates is needed to comprehensively identify genomic variations of SARS-CoV-2 in specific geographic locations.

## Data Availability

The data from the study are available in GISAID and GenBank database as mentioned in the manuscript

## Acknowledgements

We thank all the laboratory personnel involved in COVID-19 testing at the Oklahoma Animal Disease Diagnostic Laboratory. This study was funded by FDA CVM VET-LIRN.

